# Virological evidence of the impact of non-pharmaceutical interventions against COVID-19 in a resource-limited setting

**DOI:** 10.1101/2023.03.01.23286616

**Authors:** Andres Moreira-Soto, Alfredo Bruno, Doménica de Mora, Michelle Paez, Jimmy Garces, Ben Wulf, Anna-Lena Sander, Maritza Olmedo, Maria José Basantes Mantilla, Manuel Gonzalez, Alberto Orlando Nárvaez, Silvia Salgado, Juan Carlos Zevallos, Jan Felix Drexler

**Author notes:** Correspondence: Jan Felix Drexler, ^1^Charité-Universitätsmedizin Berlin, Institute of Virology, Charitéplatz 1, 10098 Berlin, Germany.

## Abstract

Ecuador was an early COVID-19 hotspot with substantial COVID-19-mortality. In developed countries, low socioeconomic status is associated with COVID-19 infection and low compliance with non-pharmaceutical interventions (NPIs). However, if NPI were successful in resource-limited settings with high human mobility and informal labour is still unclear. We performed a retrospective observational molecular and serological study of Ecuador’s reference laboratory. We tested 1,950 respiratory samples from COVID-19 surveillance for SARS-CoV-2 and 12 respiratory viruses using RT-PCR, characterized 642 SARS-CoV-2 genomes, and examined SARS-CoV-2 seroprevalence in 1,967 samples from patients with fever in Ecuador’s reference laboratory during 2020-2021. Molecular and serological data were compared to NPI stringency in Bayesian, maximum-likelihood and modelling frameworks.

SARS-CoV-2 (Pearson correlation test; r=-0.74; p=0.01) and other respiratory viruses (r=-0.68; p=0.02) detection correlated negatively with NPI stringency. SARS-CoV-2 seroprevalence increased from <1% during February-March 2020 to 50% within 6 weeks and plateaued after NPI implementation. Decrease of effective reproduction number <1 and antibody reactivity over time suggested intense SARS-CoV-2 transmission during pandemic onset, subsequently limited by NPIs. Phylogeographic analyses revealed that travel restrictions were implemented late not preventing 100 near-parallel SARS-CoV-2 introductions, and implementation of NPIs modified SARS-CoV-2 geographic spread by restricting recreational activity. NPIs stringency correlated negatively with the number of circulating SARS-CoV-2 lineages (r=-0.69; p=0.02). Virological evidence supports NPIs restricting human movement as an effective public health tool to control the spread of respiratory pathogens in resource-limited settings, providing a template for emerging SARS-CoV-2 variants and future epidemics.

## INTRODUCTION

Latin America is a COVID-19 hot spot, accumulating 20% of reported cases and 32% of deaths worldwide in 2020^1^, albeit hosting only 8% of the total world’s population. Latin America suffered substantial economic constrains due to the pandemic, exemplified by the decline of 9.4% in gross domestic product in 2020 (https://www.imf.org/en/Publications/WEO/Issues/2020/06/24/WEOUpdateJune2020). Most of the economic constraints were associated with compulsory non-pharmaceutical interventions (NPIs), i.e., policies that restrict human contact, movement, and shut down public and private services to avoid SARS-CoV-2 transmission. However, resource-limited settings such as Latin America have very high proportions of informal labour reaching more than 80% (International Labour Organization; https://ilostat.ilo.org/data/), involving high human mobility that may put the efficacy of NPIs at stake. Even in affluent settings, the efficacy of NPIs in containing COVID-19 mortality and SARS-CoV-2 spread is debated. On the one hand, early research in China and Brazil suggested that NPIs decreased SARS-CoV-2 transmission rates^2^ and reproduction number^3^, and a time series meta-analysis of 149 countries showed a decrease of COVID-19 incidence rates by 13% after NPI implementation^4^. On the other hand, a modelling study analysing European countries found no efficacy of relatively less stringent NPIs in lowering SARS-CoV-2 transmission apart from a complete lockdown^5^, and another study in Europe found that closure of businesses and stay-at-home orders were unlikely to lower COVID-19 incidence^6^.

To date, analyses of the COVID-19 public health response relied predominantly on surveillance data, leading to inevitable biases due to the limited diagnostic testing and underreporting in resource-limited settings. These biases have been partially resolved using reported excess deaths, but those data are not immediately available and depend on the quality of national data registers ^7^. Within Latin America, Ecuador was one of the earliest epicentres of the pandemic, with one of the highest COVID-19-associated mortality rates worldwide reaching 8.5% in late 2020^7^ and an 80% increase in excess deaths of baseline annual mortality^7^ despite strict and early NPI implementation. Here, we use virological data to analyse the efficacy of NPIs in Ecuador, a prototypic resource-limited setting in Latin America.

## MATERIALS AND METHODS

### Study design

Patient samples originated mainly from coastal Ecuador, separated from the rest of the country by the Andean mountains and home to most of the Ecuadorian population (**Fig. 1A**) (further geographical data shown in **Fig. S1**). That region of Ecuador was chosen because it was most affected by COVID-19 during the onset of the pandemic and because it harbours the Ecuadorian SARS-CoV-2 reference laboratory the Instituto Nacional de Investigación en Salud Pública Dr. Leopoldo Izquieta Pérez (INSPI). Other regions of Ecuador have separate and smaller reference laboratories and samples from those laboratories were hardly available for this study. We used two different subpopulations collected retrospectively from routine disease surveillance from the Ecuadorian National surveillance system (**Fig. 1B** and **1C**). First, we performed a molecular study using 1950 oro-nasopharyngeal swabs and sputum samples sent for routine SARS-CoV-2 surveillance to INSPI during March 2020 to February 2021, corresponding to 2.3% of all the samples analysed by the INSPI per week (**Fig. 1B;** further subsample data is found in **Fig. S1, S2, S3** and **S4**, and in **Table S1**). Second, all available samples of fever of unknown origin (FUO) sent to INSPI from January 2020 to February 2021 for medical investigation were used to measure SARS-CoV-2 antibodies (**Fig. 1C**; sub-population characteristics are shown in **Table S2** and **Fig. S5** and **S6**). Both subpopulations correspond to longitudinal collections of diagnostic samples spanning comparable timeframes, number of samples and geographical location (**Fig. 1D** and **1E**). Sample numbers per province were comparable between both subpopulations, and only 12% (n=243/1967) of the FUO samples were from provinces outside of coastal Ecuador from which no respiratory samples were available (**Fig. 1D** and **1E**). Although the subpopulation with FUO was younger than the subpopulation with respiratory illness, those samples are independently suitable to describe the sample reception and surveillance performed in the national reference laboratory of Ecuador during the onset of the pandemic (**Fig. S2** through **S6**).

**Figure 1.**
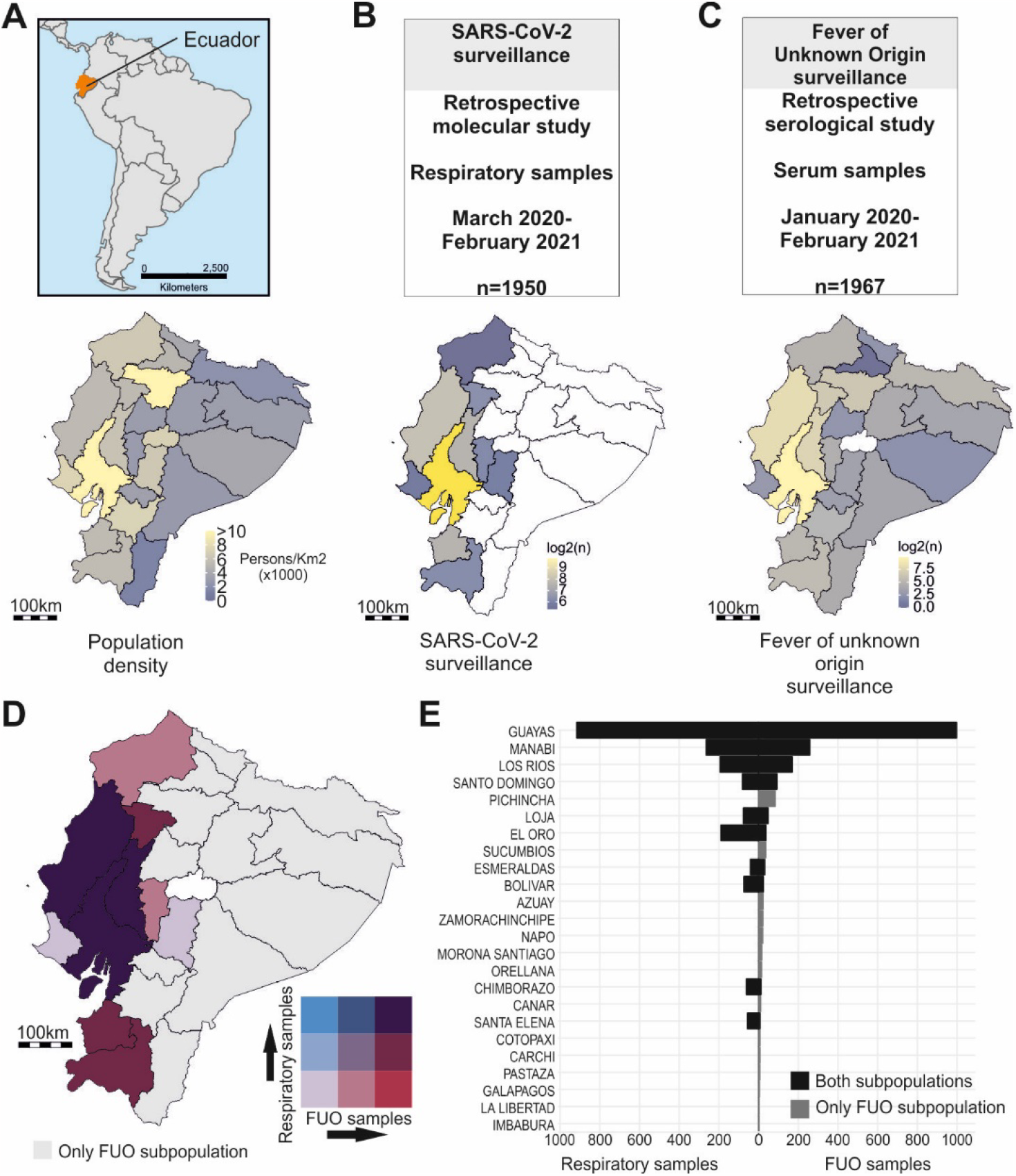
Study sites and sample collection. (A) Map of South America, Ecuador is shown in orange. Bottom: Population density map of Ecuador. (B) Subpopulation of SARS-CoV-2 surveillance used for the molecular study in Ecuador, time span of sampling, and specimens per subpopulation. Bottom: Spatial distribution of samples. (C) Subpopulation of fever of unknown origin surveillance used for the serological study in Ecuador, time span of sampling, and specimens per subpopulation. Bottom: Spatial distribution of samples. Logarithmic scale was used in (B) and (C) for clarity of presentation. (D) Bivariate map of number of respiratory and fever of unknown origin (FUO) samples in Ecuador. (E) Bar chart showing the number of respiratory samples (left) and serum samples (right) per province. Black corresponds to provinces were both subpopulations are represented. Gray corresponds to provinces where only FUO samples were available. Ecuador’s political division data (https://www.diva-gis.org/gdata; https://gadm.org/) and Ecuador’s population data (https://hub.worldpop.org/geodata/summary?id=46031; WorldPop datasets DOI: 10.5258/SOTON/WP00675) were gathered from freely available sources.

### Laboratory analyses

Molecular testing for SARS-CoV-2 and common respiratory viruses (four endemic human coronaviruses (HCoV)-OC43, -NL63, -229E, -HKU1; Human adenoviruses, metapneumovirus, parechovirus, influenza A/B virus, respiratory syncytial viruses A/B, enteroviruses and parainfluenza viruses 1-4) was done using commercially available multiplex real-time RT-PCR kits, SARS-CoV-2 whole-genome amplification and assembly using readily available pipelines (https://gitlab.com/RKIBioinformaticsPipelines/ncov_minipipe) and classification using (PANGOLIN) version 3.1.16^8^. Serological testing included a validated SARS-CoV-2 testing algorithm based on a chemiluminescence immunoassay followed by a highly specific SARS-CoV-2 surrogate virus neutralization test described before^9^.

### Evolutionary analyses

We constructed an approximately-maximum likelihood (ML) phylogeny encompassing all SARS-CoV-2 sequences from this study and all available GISAID genomes until September 1, 2021 using the program fasttree 2.1.10^10^ with a GTR+CAT substitution model. To explore the temporal signal of SARS-CoV-2, an ancestral state inference in an ML framework was calculated in TreeTime 0.7.6^11^. Phylogeographic inference was calculated using the relaxed random walk (RRW) diffusion model run for 1□×□10^8^ generations implemented in BEAST v1.10.4.

### Statistical analyses and modelling

Chi-square tests of proportions and Fisher’s exact tests were used to compare categorical variables, t-tests and Pearson correlation tests to compare continuous variables using R (v. 4.0.3). Kernel density estimations (KDE), used to visualize the distribution of continuous variables in time were performed in R (v. 4.0.3). KDE were calculated using the smallest bandwidth possible due to our large sample size. COVID-19 outbreak dynamics were analysed using a susceptible-exposed-infectious-recovered (SEIR) model modified to fit COVID-19 estimates^12^. Further details on materials and methods are found in the **Supplementary text**.

## RESULTS

### NPI implementation in Ecuador

Ecuador’s public health response to COVID-19 started immediately after declaration of the pandemic by the WHO on March 11, 2020^13^. Ecuador’s government implemented an emergency state, handing pandemic response to the central government. The implemented NPIs increased in severity and included closing of national borders on March 16, followed by banning social gatherings, a curfew, closing of public spaces, commerce, schools, and stay-at-home orders by March 23, corresponding to a high NPI stringency level of 93% according to the widely used Oxford-based classification system^14^. The Stringency index is calculated as the mean score of nine metrics: school closures; workplace closures; cancellation of public events; restrictions on public gatherings; closures of public transport; stay-at-home requirements; public information campaigns; restrictions on internal movements; and international travel controls. In Ecuador, the NPI stringency index decreased 3-7% per month until mid-September 2020, time in which Ecuador central government lifted stay-at-home orders, travel restrictions, permitted public gatherings, and gave political power to local governments to decide on NPIs. Afterwards, the stringency index was approximately 50% for the rest of the year (stringency of NPI over time is shown in **Fig. S10**). Of note, the obligatory use of masks in indoor and outdoor spaces was implemented on April 6, 2020 and lifted on April 2022.

### NPIs likely constrained circulation of SARS-CoV-2 and other respiratory viruses

During the onset of the pandemic, Ecuador’s laboratory testing capacity was below all other South American countries, reaching 40 tests per 1000 inhabitants as of December, 2020^1^, and lack of reagents challenged homogenous testing over time (total number of tests shown in grey in **Fig. 2A**) (https://www.theguardian.com/world/live/2020/jun/12/coronavirus-live-news-markets-fall-over-fears-of-long-us-recovery-as-brazil-cases-top-800000). Therefore, we re-tested 1,950 samples stored at Ecuador’s reference laboratory continuously during the first year of the COVID-19 pandemic (mean, 47.8 samples per week (range: 27.5 to 60). In total, 52% (1017/1950) of those samples were SARS-CoV-2 positive. The high RT-PCR detection rate is consistent with the Ecuadorian testing algorithm focusing on symptomatic cases and contacts ^15^. The SARS-CoV-2 RT-PCR detection per month correlated significantly with lower stringency of NPIs, implying increased virus circulation before NPIs were implemented and after NPIs were relaxed (Pearson correlation test; r=-0.74; *p*=0.01) (**Fig. 2B**). The positivity rate decreased slowly with time, from 40% in March 2020, when the more stringent NPIs were implemented, until 20% in September 2020, were NPIs were the lowest, suggesting that NPIs were successful in controlling virus spread. As soon as NPI stringency was lowered, the positivity rate got a steep increase reaching close to 40% during late 2020, following the reported excess mortality increase (**Fig. 2A**).

**Figure 2.**
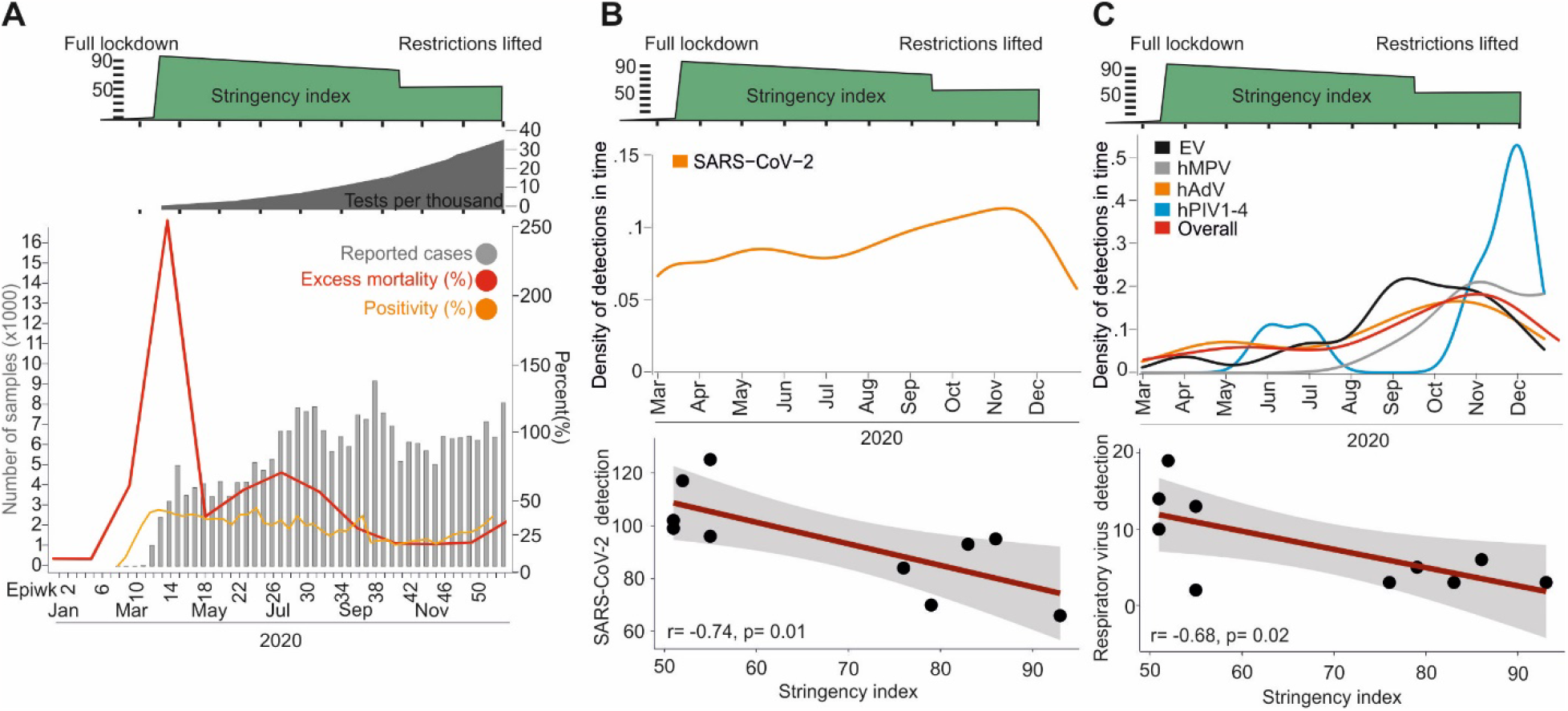
Circulation of SARS-CoV-2 and other respiratory viruses in Ecuador. (A) Reported number of cases and all-cause excess mortality from the Ecuadorian Ministry of Health (www.salud.gob.ec) and positivity rate from Ecuador’s reference Laboratory. Above: Total number of tests per thousand inhabitants (Our World in Data) and Oxford stringency index. (B) Kernel density estimations of SARS-CoV-2 detections. Below: Pearson correlation test of NPI stringency index versus number of SARS-CoV-2 detections per month. (C) Kernel density estimations of non-SARS-CoV-2 respiratory viruses. Below: Pearson correlation test of NPI stringency index versus number of respiratory virus detections per month. Viruses with less than two detections are not shown for graphical reasons. Human adenoviruses (hAdV), metapneumovirus (hMPV), enteroviruses (EV) and parainfluenza viruses 1-4 (hPIV 1-4). Epiwk: Epidemiological week.

The overall detection rate of all other respiratory viruses was low at 4.7% (91/1950; CI: 3.9-5.8) (**Fig. 2C**). Among common respiratory viruses, adenoviruses (Fisher’s exact-test; p=0.026), parainfluenzaviruses (Fisher’s exact-test; p=0.04), enteroviruses (Fisher’s exact-test; p<0.0001) and metapneumoviruses (Fisher’s exact-test; p<0.0001) occurred significantly more frequently during months of non-stringent NPIs, being the odds of detecting respiratory viruses 2-23 times higher in lower NPI as in stringent NPIs times (individual detection of respiratory viruses in SARS-CoV-2-positive and -negative patients are shown in **Fig. S13**). Similar to SARS-CoV-2, the detection of those common respiratory viruses significantly correlated with less stringent NPIs (Pearson correlation test; r=-0.68; *p*=0.02) (**Fig. 2C**), reminiscent of the reduced circulation of SARS-CoV-2 and suggesting modified respiratory virus circulation according to NPI stringency. Therefore, there was reduced respiratory virus circulation in comparison to pre-pandemic detection rates in Ecuador^16^. Co-infection between SARS-CoV-2 and other respiratory viruses was detected in 4.4% of SARS-CoV-2 positive samples (44/1017; CI: 6.1-9.4), and only in one other sample by non-SARS-CoV-2 respiratory viruses (**Fig. S13**). The detected rates of co-infection were considerably lower than co-infection rates found in pre-pandemic studies from tropical settings, reaching more than 10% using similar methodology^17^. Overall decreased virus circulation was likely associated with stringent NPIs and included the complete absence of influenza viruses, consistent with a study from the United Kingdom^18^. Finally, co-infections did not affect COVID-19 severity or duration (data on disease outcome are found in **Fig. S14)**.

### SARS-CoV-2 seroprevalence plateaued following the implementation of NPIs

SARS-CoV-2-specific IgG antibodies were first detected during late March 2020 (**Fig. 3A**). IgG seroconversion occurs in patients approximately after the third week of illness^19^. Therefore, the serological data was in concordance with the first SARS-CoV-2 case detected in the country on February 29^th^. One month later, during early May 2020, SARS-CoV-2 seroprevalence reached 50% and stagnated afterwards (**Fig. 3A**), compatible with intense SARS-CoV-2 transmission during the onset of the pandemic mainly in coastal Ecuador, followed by limited onward transmission due to NPIs (**Fig. S7, S8** and **S9**). This interpretation was consistent with decreasing levels of antibodies over time in our dataset (**Fig. 3B**; **Fig. S7**) and the modelled effective reproduction number (R_eff_) <1 using seroprevalence and incidence data, during stringent NPIs (**Fig. 3C**), and reminiscent of an observational study showing decreased SARS-CoV-2 circulation due to implementation of NPIs in Hong Kong^20^ and a modelled decrease of R_eff_ estimates due to NPIs from São Paulo and Rio de Janeiro, Brazil during the onset of the pandemic ^3^. Given the relatively young age of patients in this subpopulation, the overall seroprevalence in Ecuador may be under or overestimated. However, comparable seroprevalence estimates in a population-based study from Peru showed an equal seroprevalence distribution amongst age groups^21^. From September 2020 onwards, R_eff_ re-increased >1, and from December 2020 onwards, mean CLIA antibody reactivity levels re-increased, likely due to relaxed NPIs leading to increased virus transmission (**Fig. 3C** and **Fig. S7**).

**Figure 3.**
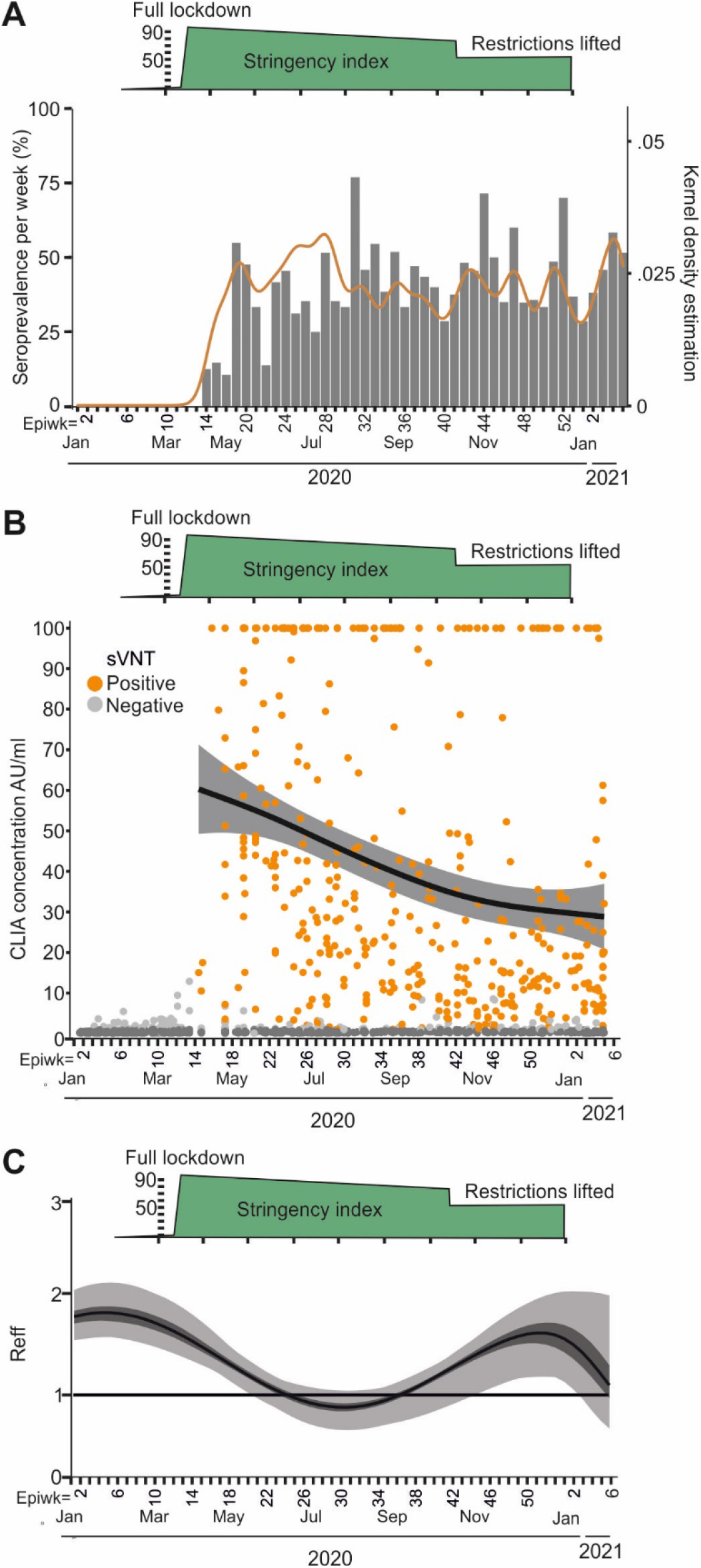
Seroprevalence and transmission of SARS-CoV-2 in Ecuador. (A) Seroprevalence per epidemiological week. Right axis denotes kernel density estimation analyses of SARS-CoV-2-specific antibodies in Ecuador. (B) CLIA concentration across time in the tested samples. The black line denotes the mean CLIA concentration per month. Gray lines denote the 95% confidence interval. AU/ml: absorbance units per millilitre. (C) Calculated effective reproduction number from seroprevalence estimates and reported cases over time. Epiwk: Epidemiological week.

### Travel restrictions were implemented too late to prevent parallel SARS-CoV-2 introductions

Since there is a lack of SARS-CoV-2 genomic data from the onset of the COVID-19 pandemic in Ecuador^22^, we sequenced 63.1% (642/1017) of the SARS-CoV-2-positive samples. Phylogenomic reconstructions combining our new data and all SARS-CoV-2 sequences from Ecuador in GISAID until September 2021 revealed that SARS-CoV-2 was introduced into Ecuador between February 17 to March 8, 2020 (**Fig. 4A**). The time of introduction was similar to estimates from Brazil, the first country in Latin America detecting SARS-CoV-2 ^3^, and thus compatible with early spread of COVID-19 in Ecuador and robustness of our data. That time span also included the first reported case of SARS-CoV-2 infection from Ecuador and was in concordance with our serological data, again suggesting robustness of our results. Close to 100 separate SARS-CoV-2 introduction events into Ecuador were suggested by the low average genetic divergence, the similar detection time and the clustering of the early SARS-CoV-2 viruses in the time stamped tree in early SARS-CoV-2 sequences from Ecuador (**Fig. 4A**; see divergence data in **Fig. S15** and **S16**). Although Ecuador was among the first Latin American countries to implement travel restrictions by March 16, 2020, those NPIs were apparently implemented too late to prevent multiple parallel introduction events. For comparison, similarly high numbers of parallel SARS-CoV-2 introductions were reported during the onset of COVID-19 in Brazil, again suggesting robustness and generalizability of our data ^3^.

**Figure 4.**
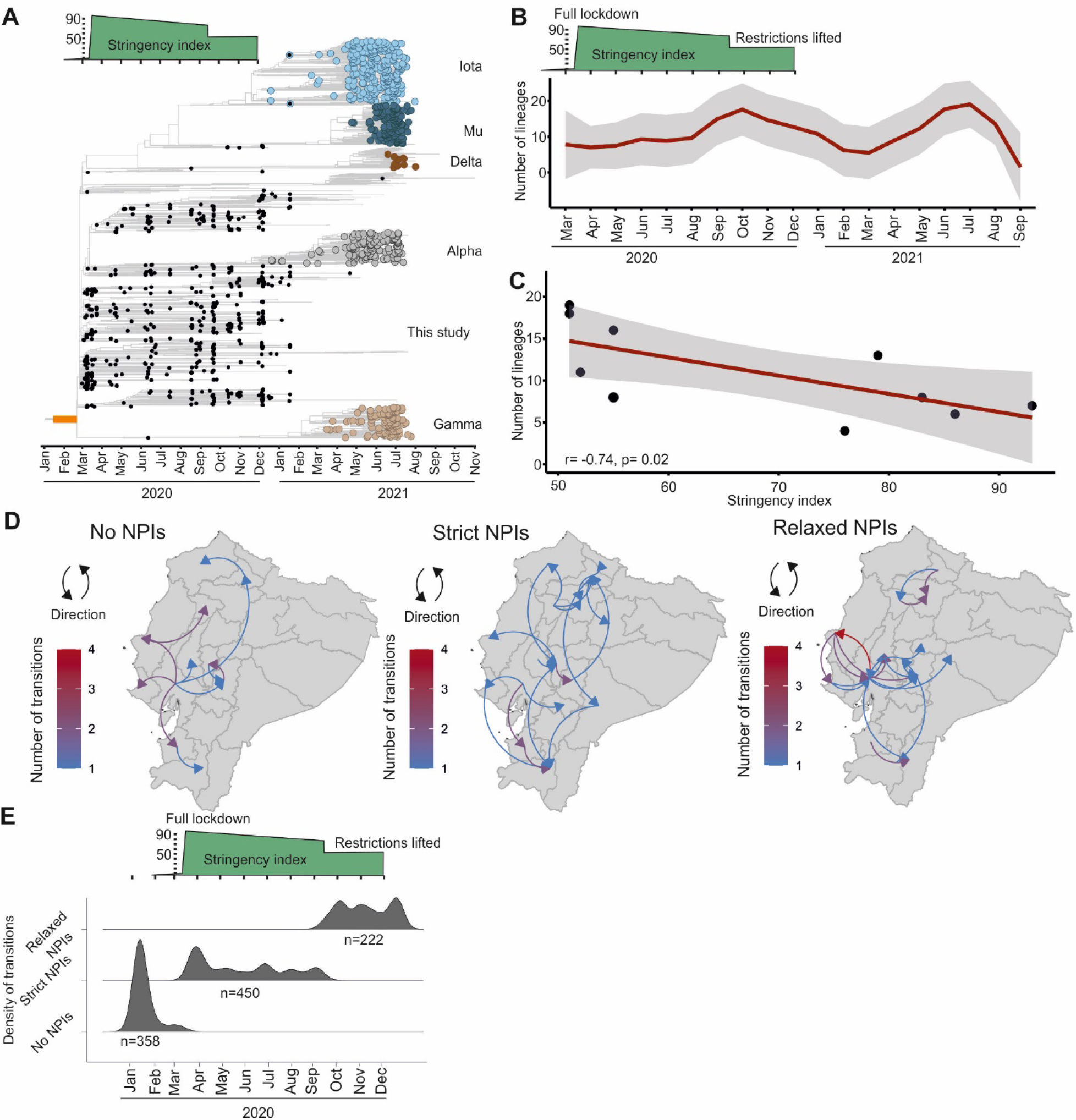
Evolution and spread of SARS-CoV-2 in Ecuador. (A) Time-resolved maximum clade credibility phylogeny of SARS-CoV-2 in Ecuador. Sequences generated for this study in black. Variants of concern or of interest are coloured. The time of SARS-CoV-2 introduction is shown in an orange square at the root of the tree. (B) Number of distinct Pango lineages in time. Gray zone represents 95% confidence intervals. (C) Pearson correlation test of stringency index versus number of lineages per month. Gray zone represents 95% confidence intervals. (D) Spatiotemporal reconstruction of the spread of SARS-CoV-2 in Ecuador during establishment of non-pharmaceutical interventions (NPIs). NPI implementation time as in (D). (E) Density estimations of SARS-CoV-2 transitions.

### NPI implementation slowed the emergence of new SARS-CoV-2 lineages

The SARS-CoV-2 strains circulating in Ecuador during 2020 and 2021 were assigned to 66 different lineages (see distribution of Pango lineages over time in **Fig. S17** and **S18**). The early phase of the pandemic in Ecuador was characterized by the predominance of lineages B.1, B.1.1 and its sub-lineages; B.1.1 representing 30% of the total genomic data. Both B.1 and B.1.1. lineages were detected during February 2020 in Europe and to a lesser extent in North America and Asia ^23^, and in Ecuador during March 2020, again compatible with travel-aided SARS-CoV-2 introduction events into Ecuador. In our dataset, a mean of 7.6 (range: 4-13; CI: 4.4-10.8) SARS-CoV-2 Pango lineages were detected during March to August 2020. Once NPIs were relaxed from September to December 2020, almost twice as many lineages circulated despite the shorter time span (mean, 16; range: 11-19; CI: 10.4-21.6) (**Fig. 4B)**. Moreover, the number of lineages was negatively correlated with the stringency of NPIs, suggesting accelerated viral evolution due to increased transmission and introduction of previously non-endemic lineages once NPIs were relaxed (**Fig. 4C**) (Pearson correlation test; (r=-0.69; p=0.02).

### NPIs modified the dispersion of SARS-CoV-2

Phylogeographic reconstructions of the regionally predominant SARS-CoV-2 lineage B.1.1 showed differential dispersion patterns during different phases of the pandemic. At the onset of the pandemic, geographic transitions originated unidirectionally from coastal Ecuador to other provinces (**Fig. 4D**), which was in concordance to our time-stamped analyses, and in concordance to the natural progression of SARS-CoV-2 geographic spread. Two weeks after the implementation of strict NPIs, a 54% decrease of geographic transitions entailed, followed by continuously low numbers of geographic transitions (**Fig. 4D** and **4E**). Virus movements during strict NPIs occurred multidirectionally over relatively larger distances (**Fig. 4D**), potentially due to increased mobility associated with commercial reactivation during mid-2020 in all of South America ^24^, since travel restrictions and stay-at-home orders were still implemented. Once NPIs were relaxed, transitions peaked again and occurred predominantly over shorter distances, potentially associated with recreational activity, as a high number of transitions occurred to and from coastal provinces (**Fig. 4D** and **4E**).

## DISCUSSION

We gathered virological data describing the onset of the COVID-19 pandemic in a Latin American COVID-19 epicentre to show that NPIs were a modifying and limiting factor of SARS-CoV-2 transmission. NPIs are a well-known part of public health responses to epidemics and have been used, among others, during the SARS-CoV epidemic in 2003 and the Spanish flu in 1918. However, the success of NPIs in containing outbreaks varied depending on several factors, including region, time of implementation, cultural background of the population, its prior experience to epidemics and civil compliance to the imposed restrictions ^25,26^. For SARS-CoV-2, a comparative study analysing public health responses in developed and a limited number of developing countries suggested that the success of NPIs depended on governmental monitoring and early and stringent application of NPIs ^27^, which are particularly difficult in resource-limited settings such as Latin America due to several factors. First, testing of SARS-CoV-2 in Latin America is insufficient and assessments of base mortality are often delayed ^7^. Second, monitoring and implementation of NPIs is complicated in rural settings, in which circa 20% of Latin American populations reside ^9^ and in which access to healthcare services is limited and COVID-19 stigmatization, i.e. delay of seeking care due to negative economic or social consequences, is frequent ^9,28^. Third, compliance with NPIs by the population is affected by socio-demographic characteristics such as poverty and ethnicity ^29^ and Latin American societies are prototypic examples of resource-limited multi-ethnic settings. Fourth and potentially most importantly, informal labor reaches more than 60% of workforce in Ecuador and other parts of Latin America ^30^, and stay-at-home orders cannot be followed by persons relying on daily informal income and lacking infrastructure to store food. Those factors may explain why despite strict NPIs, the death toll continued to rise in Ecuador and other regions of Latin America, and why Latin America is a priority region for rapid vaccination ^31^. Whether NPIs were thus a meaningful tool to contain COVID-19 in Latin America and whether the socio-economic costs of NPIs were justified thus remained unclear ^32^. In stark contrast, our data reveal an impact of NPIs on all aspects of SARS-CoV-2 epidemiology and strongly suggest that without NPIs, the death toll would have been much higher during the first year of the pandemic in Latin America, reminiscent of a retrospective modelling study on NPI implementation in 2020 in India, which projected a 60% reduction in deaths compared to non-implementation of NPIs ^33^.

### Limitations

Limitations of our study include the use of serum and respiratory samples that were not designed to represent Ecuador’s population and that we assume no reduction in compliance over time of NPIs, which are less followed by persons with low socioeconomic status ^34^. Additionally, the use of the Oxford stringency index summarizing NPIs considers measures isolated from regional contexts, such as informal labour proportion and the degree of enforcement of NPIs by regional authorities. However, using policy response aggregates might be of use because individual policies during the first pandemic year had limited variation across and within countries, resulting in collinear relationships ^14^. Another limitation are the intrinsic biases of phylogenetic analyses due to representability of spatiotemporal samples and the lack of a bigger genomic dataset from Ecuador in comparison to other Latin American countries such as Brazil ^3^. However, our samples are representative of the Ecuadorian national testing algorithm and taken at even rates continuously across the first year of the COVID-19 pandemic and should thus allow robust interpretations despite inevitable sample biases, consistent with similar detection patterns of SARS-CoV-2 and other respiratory viruses. Phylogenomic analyses from an affluent setting confirmed usability of genomic data to analyse NPI efficacy ^35^. Additionally, acute febrile disease from arbovirus infections or malaria is common in poorer population strata ^12^ and those sera should allow sensitive detection of past SARS-CoV-2 infection which is also common in poorer population strata ^36^.

Methodological strengths of our study include the use of exhaustive molecular, phylogenetic, modelling and serological analyses whose results mutually confirmed each other and compensate for the study’s inherent limitations.

## Conclusion

Our data support that stringent NPIs modifying human behaviour, e.g., limiting human movement, social gatherings and increasing available information to the public were efficient against COVID-19 in Ecuador, but intense SARS-CoV-2 spread occurred before their implementation. Therefore, NPIs should be rapidly implemented and sustained in immunologically naïve populations upon introduction of a highly transmissible pathogen such as SARS-CoV-2. This interpretation is in concordance with studies showing increased mortality when NPIs were introduced too late and lifted too early during the 1918 Spanish flu pandemic in the U.S. ^26^ and from early travel restrictions globally reducing case numbers during the onset of the COVID-19 pandemic ^37^. Finally, the emergence of antigenically distinct SARS-CoV-2 variants worldwide ^38^ leading to decreased vaccine effectiveness ^39^, together with the delayed modification of vaccines worldwide has led countries such as China, Canada and Peru to implement similarly stringent NPIs as in 2020, affecting hundreds of millions of people in 2022. Overall, our analyses of virological data provide an evidence-based way to justify the implementation of NPIs, providing data-driven support to stakeholders facing resurge of SARS-CoV-2 immune escape variants or future epidemics globally. Nonetheless, the virological benefit of NPIs must be weighed against the capacities of public health systems in managing severe COVID-19 cases, e.g. intensive care units, respirators, and oxygen, all limited in resource-limited settings like Latin America ^40,41^ and the high societal ^42^ and economic costs associated with NPIs.

## Data Availability

All data needed to evaluate the conclusions in the paper are present in the paper and/or the Supplementary materials. The data that is not patient associated will be made available for research on reasonable request and with permission from the INSPI and the Ecuadorian ministry of health. Newly generated SARS-CoV-2 sequences will be uploaded to GISAID. The SARS-CoV-2 sequence assembly can be found in ebi.ac.uk under the accession: ERP145042. All the code to reproduce the analyses can be found at: https://github.com/Dokandres/EcuadorNPI

## Acknowledgments

We thank Arne Kühne for technical support. We thank Carlo Fischer for the critical reading of the manuscript.

## Ethical statement

All patient data were managed in anonymized databases by the INSPI. Clinical symptoms were retrieved from medical charts. The study was approved by the Ecuadorian Ministry of Health (MSP-MSP-2021-0006-O-FDQ) and the Ethics committee of the Espiritu Santo University in Guayaquil, Ecuador (2022-002A).

## Author contributions

Conceptualization (AM-S, JCZ, JFD), data curation (AB, DdM, MP, JG,JG, BW, A-LS, MO, MJB, MG, AO, SS), formal analysis (AM-S, AB, DdM, MP, JG, BW, A-LS, MO, MJB, MG, AO, SS, JCZ, JFD), funding acquisition (JFD), investigation (AM-S, AB, DdM, MP, JG, BW, A-LS, MO, MJB, MG, AO, SS, JCZ, JFD), methodology (AM-S, AB, DdM, MP, JG, BW, A-LS, MO, MJB, MG, AO, SS, JCZ, JFD), project administration (AM-S, AO, JCZ, JFD), resources (AM-S, AB, DdM, MP, JG, BW, A-LS, MO, MJB, MG, AO, SS, JCZ, JFD), software (AM-S, A-LS, BW), visualization (AM-S, BW, A-LS), writing – original draft (AM-S, AB, DdM, MP, JG, BW, A-LS, MO, MJB, MG, AO, SS, JCZ, JFD), and writing – review & editing (AM-S, JFD)

## Data and materials availability

All data needed to evaluate the conclusions in the paper are present in the paper and/or the Supplementary materials. The data that is not patient associated will be made available for research on reasonable request and with permission from the INSPI and the Ecuadorian ministry of health. Newly generated SARS-CoV-2 sequences were uploaded to GISAID (numbers: XXXXXXXXXXX-XXXXXXXXXXX). The SARS-CoV-2 sequence assembly can be found in ebi.ac.uk under the accession: ERP145042. All the code to reproduce the analyses can be found at: https://github.com/Dokandres/EcuadorNPI.

