## Supplementary methods and figures for "Virological evidence of the impact of non-pharmaceutical interventions against COVID-19 in a resource-limited setting"

Correspondence:

**Supplementary text. Materials and methods.**

**Subpopulation characteristics:** For the molecular study, a total of 1950 oro-nasopharyngeal swabs and sputum samples were used. The median age of the subpopulation was 40 years (interquartile range: 29-55), corresponding to 48.5 % (946) female and 51.5% (1004) male patients (**Fig. S3** and **Table S1**). The number of samples gathered per province was comparable with the total population (**Fig. S2**). For COVID-19 serology, 1967 sera from patients with fever of unknown origin (FUO) sent to INSPI from January 2020 to February 2021. Further sample data are found in **Fig. S1, S2, S3; S4** and **S5,** and in **Table S1** and **S2**) A total of 1967 samples were gathered from 22 out of the 24 provinces of Ecuador. The median age within this subpopulation was 14 years (interquartile range: 8-27), corresponding to 49% (947) female and 51% (970) male patients (**Fig. S5** and **Table S2).**

**Molecular testing:** We performed nucleic acid purification using the MagNA Pure 96 DNA and Viral NA small-volume kit following the manufacturer's instructions. (Roche, Penzberg, Germany). SARS-CoV-2 testing was performed using the SarbecoV E-gene and RdRP real time RT-PCR-based kits (TIB Molbiol, Germany). Multiplexed testing for common respiratory viruses was performed using commercially available multiplex real-time RT-PCR kits (TIB Molbiol) targeting the four endemic human coronaviruses (HCoV)-OC43, -NL63, -229E, -HKU1; Human adenoviruses, metapneumovirus, parechovirus, influenza A/B virus, respiratory syncytial viruses A/B, enteroviruses and parainfluenza viruses 1-4.

**Genomic characterization:** Whole-genome amplification of SARS-CoV-2 was done using the Artic V3 PCR-based protocol (https://artic.network/ncov-2019). Library preparation was done using the KAPA Frag kit and KAPA Hyper Prep kit (Roche Molecular Diagnostics, Switzerland) and sequencing was done using MiSeq reagent v2 chemistry (Illumina, USA) according to the manufacturers' protocols. Genome assembly was done by mapping MiSeq reads to the Wuhan-Hu-1 SARS-CoV-2 reference sequence (GenBank accession number: NC045512) using the Python-based CoVpipe pipeline (https://gitlab.com/RKIBioinformaticsPipelines/ncov_minipipe). COVID-19 lineage assignment was performed using a dynamic lineage classification method called Phylogenetic Assignment of Named Global Outbreak LINeages (PANGOLIN) version 3.1.16 (1).

**Serologic testing:** Dengue is endemic in coastal Ecuador. Nevertheless, the years 2020 and 2021 saw a two-fold increase in Dengue cases compared to 2019, suggesting that both DENV and SARS-CoV-2 were circulating at high rates in the population (https://www3.paho.org/data/index.php/es/temas/indicadores-dengue/) (**Fig. S6**). Accordingly, 50% (n=1538/1967) of the FUO samples were tested for acute Dengue virus (DENV) infection targeting the NS1 protein and IgM antibodies in an enzyme-linked immunosorbent assay (J. Mitra, India) (**Fig. S6**). Of the tested samples, circa 48% (742/1538) were positive for one or both DENV-specific assays, confirming a high DENV circulation in Ecuador. Due to unspecific reactivity elicited by endemic tropical diseases such as malaria, Dengue, and Zika (2, 3), we used a validated two-step testing algorithm to ensure robust serologic results for SARS-CoV-2 IgG antibodies (4). Briefly, screening for SARS-CoV-2-specific IgG antibodies was done using a chemiluminescence immunoassay using the SARS-CoV-2 spike receptor-binding domain as antigen (CLIA; SARS-CoV-2 S-RBD IgG kit; Snibe Diagnostic, China) followed by a highly specific SARS-CoV-2 surrogate virus neutralization test (sVNT; GenScript, USA) in all CLIA-reactive samples. Only samples yielding positive results in both SARS-CoV-2 assays were considered for further analyses (4) (assay data can be found in **Fig. S7**). Since seroprevalence estimates were comparable irrespective of whether samples outside of coastal Ecuador were included, we performed further analyses using the whole FUO subpopulation (**Fig. S8 and Fig. S9)**. Ecuador started vaccination against COVID-19 in January 2021. By February 2021, when last serum samples used in this study were sampled, there were 0.43 vaccines administered per 100 persons, precluding bias of our serological analyses by vaccine-associated immune responses (5).

**Evolutionary analyses:** Sequences were aligned using mafft v7.445 (6). An approximately-maximum likelihood (ML) phylogeny encompassing all SARS-CoV-2 sequences from this study and all available GISAID genomes until September 1, 2021 was reconstructed using the program fasttree 2.1.10 (7) with a GTR+CAT substitution model. To explore the temporal signal of SARS-CoV-2, an ancestral state inference in an ML framework was calculated in TreeTime 0.7.6 (8), using a fixed clock rate calculated previously for SARS-CoV-2 of 8×10^-4 substitutions per site per year and a standard deviation of 4×10^-4 (https://docs.nextstrain.org/projects/ncov/en/latest/reference/configuration.html; Nextstrain SARS-CoV-2 Workflow development version:8fdf1932).

Phylogeographic inference was calculated using the relaxed random walk (RRW) diffusion model run for $1 \times 10^8$ generations implemented in BEAST v1.10.4 (9), using a dataset comprising SARS-CoV-2 B.1.1 Pango lineage sequences only to limit biases from different transmission dynamics between lineages; and because B.1.1 was the only lineage detected throughout the whole study period. The R package "seraphim" (10) was used to extract spatio-temporal information in the posterior trees dataset and visualize the continuous phylogeographic reconstructions using the discrete variables: No-NPI, Strict-NPI and relaxed-NPI.

**Susceptible-exposed-infectious-recovered (SEIR) model:** The model was implemented in a Bayesian framework using the LibBi library via the RBi and RBi.helpers packages. The model was jointly fitted to reported COVID-19 incidence data from Ecuador gathered from Our World in Data (11), from the first case onwards until February 28th, 2021 and the proportion of individuals who were seropositive to COVID-19 after the stabilization of the seroprevalence observed in the distribution of kernel density estimation analyses from the epidemiological week 19 (March 19, 2020) onwards until epidemiological week 5 (February 6, 2021). Incidence was fitted using a Poisson likelihood with overdispersion and approximated with a truncated Gaussian distribution, and seroprevalence was fitted using a binomial likelihood. Informative prior probability distributions were used for the delay between infection and infectiousness centred around 3,6 (standard deviation (SD)=0.7 days) (12). Additionally, replication-competent virus cannot usually be recovered from individuals with mild-to-moderate COVID-19 disease beyond 10 days of symptom onset, therefore we centred the infectious period around 10 days (SD=2.4) using a lognormal distribution (13, 14). Uniform prior probability distributions were used for proportion of cases reported, and basic reproduction number. Regularizing prior probability distributions were used for the initial numbers of infected and overdispersion of reporting. All model parameters were estimated using Markov chain Monte Carlo (full prior and posterior distributions are found in **Fig. S11** and S**12**). Further model data was described in (15).

**Statistical analyses and modelling:** Georeferencing was performed in R (v. 4.0.3) using open source maps of Ecuador´s political division (https://www.diva-gis.org/gdata; https://gadm.org/) and Ecuador´s population (https://hub.worldpop.org/geodata/summary?id=46031; WorldPop datasets DOI : 10.5258/SOTON/WP00675) . The OxCGRT stringency index was gathered from Our World in Data (11) (**Fig. S10**).

All the code to reproduce the analyses can be found at:
https://github.com/Dokandres/EcuadorNPI

1. Rambaut A, Holmes EC, O'Toole Á, Hill V, McCrone JT, Ruis C, et al. A dynamic nomenclature proposal for SARS-CoV-2 lineages to assist genomic epidemiology. Nature Microbiology. 2020 2020/11/01;5(11):1403-7.
2. Yadouleton A, Sander AL, Moreira-Soto A, Tchibozo C, Hounkanrin G, Badou Y, et al. Limited Specificity of Serologic Tests for SARS-CoV-2 Antibody Detection, Benin. Emerg Infect Dis. 2021 Jan;27(1).
3. Lustig Y, Keler S, Kolodny R, Ben-Tal N, Atias-Varon D, Shlush E, et al. Potential Antigenic Cross-reactivity Between Severe Acute Respiratory Syndrome Coronavirus 2 (SARS-CoV-2) and Dengue Viruses. Clinical infectious diseases : an official publication of the Infectious Diseases Society of America. 2021 Oct 5;73(7):e2444-e9.
4. Moreira-Soto A, Pachamora Diaz JM, Gonzalez-Auza L, Merino Merino XJ, Schwalb A, Drosten C, et al. High SARS-CoV-2 Seroprevalence in Rural Peru, 2021: a Cross-Sectional Population-Based Study. mSphere. 2021 Dec 22;6(6):e0068521.
5. Roser M. Coronavirus Pandemic (COVID-19). Our World in Data. 2020 2020.
6. Katoh K, Standley DM. MAFFT Multiple Sequence Alignment Software Version 7: Improvements in Performance and Usability. Molecular Biology and Evolution. 2013;30(4):772-80.
7. Price MN, Dehal PS, Arkin AP. FastTree 2--approximately maximum-likelihood trees for large alignments. PLoS One. 2010 Mar 10;5(3):e9490.
8. Sagulenko P, Puller V, Neher RA. TreeTime: Maximum-likelihood phylodynamic analysis. Virus Evol. 2018 Jan;4(1):vex042.
9. Suchard MA, Lemey P, Baele G, Ayres DL, Drummond AJ, Rambaut A. Bayesian phylogenetic and phylodynamic data integration using BEAST 1.10. Virus Evol. 2018 Jan;4(1):vey016.

**Section 1. Sample geographical data**

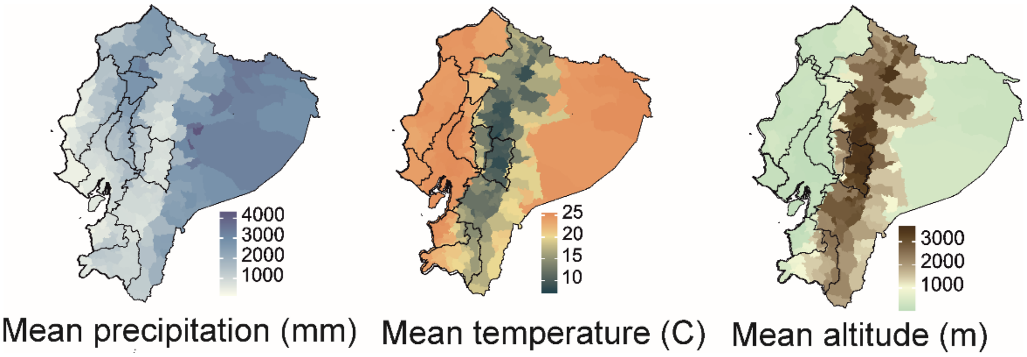

**Figure S1. Ecuador mean precipitation (millimeter), mean temperature (Celsius), and mean altitude (meter) maps in the provinces from which the respiratory samples were mostly gathered.**

The regions sampled are marked in black. Data gathered from https://www.worldclim.org/.

**Section 2: Sample characteristics**

***Section 2.1: Characteristics of the respiratory samples sent to the National Institute of Public Health Research (INSPI), Guayaquil, Ecuador, 2020-2021***

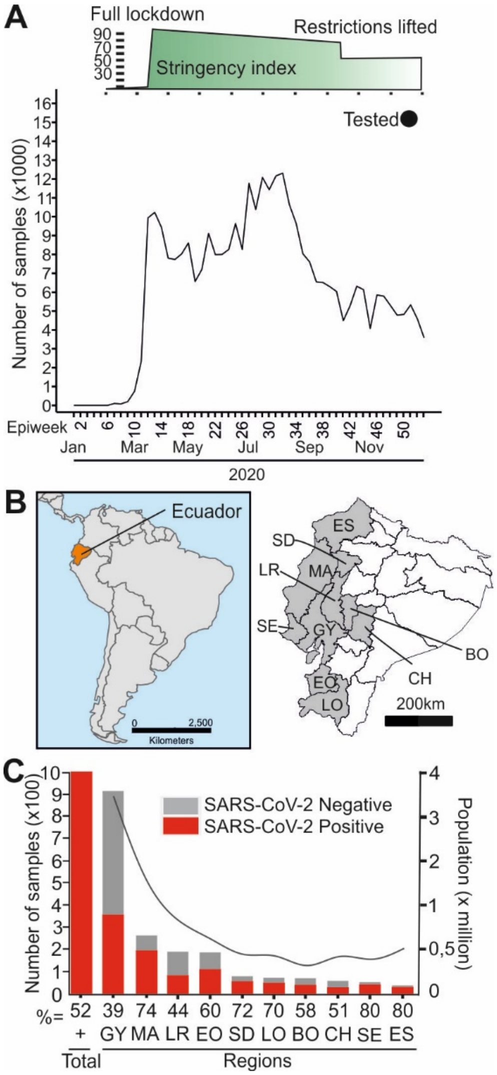

**Figure S2. Number of samples received by the reference laboratory (INSPI).**

(A) Number of tested samples in Ecuador's reference laboratory. (B) Map of South America, Ecuador is shown in orange. Right: map of Ecuador highlighting the regions sampled in gray. ES= Esmeraldas, MA= Manta, SD= Santo Domingo de los Colorados,

LR= Los Rios, SE= Santa Elena, GY= Guayas, BO= Bolivar, CH=Chimborazo, EO= El Oro LO= Loja. (C) Sample number (left axis) and population in millions (right axis) of the regions sampled. Abbreviations as in. Population data gathered from https://www.citypopulation.de/en/ecuador/cities/. The number of samples per province is in concordance with the total population per province.

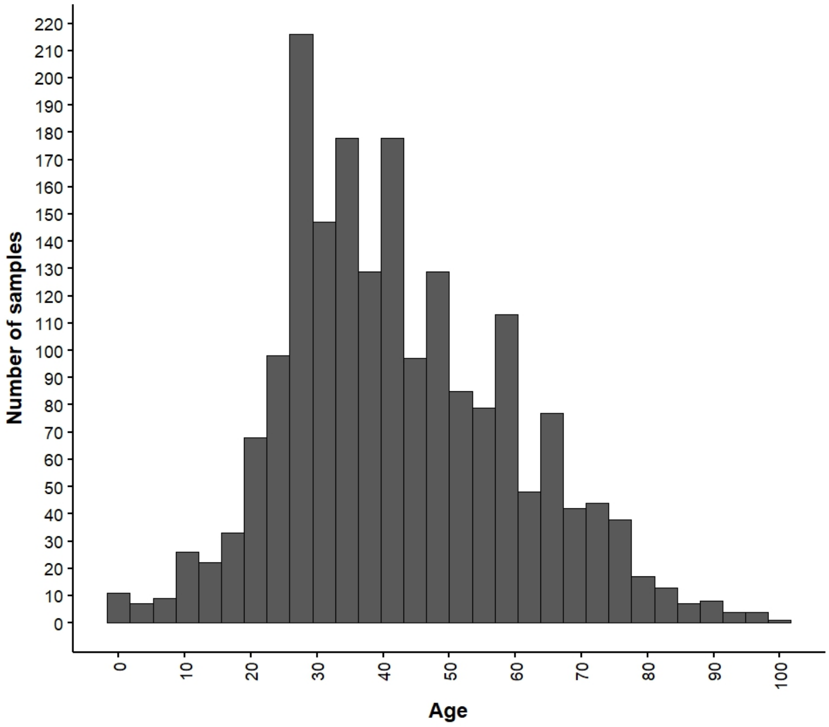

**Figure S3. Age distribution of the patients for which respiratory samples were available.**

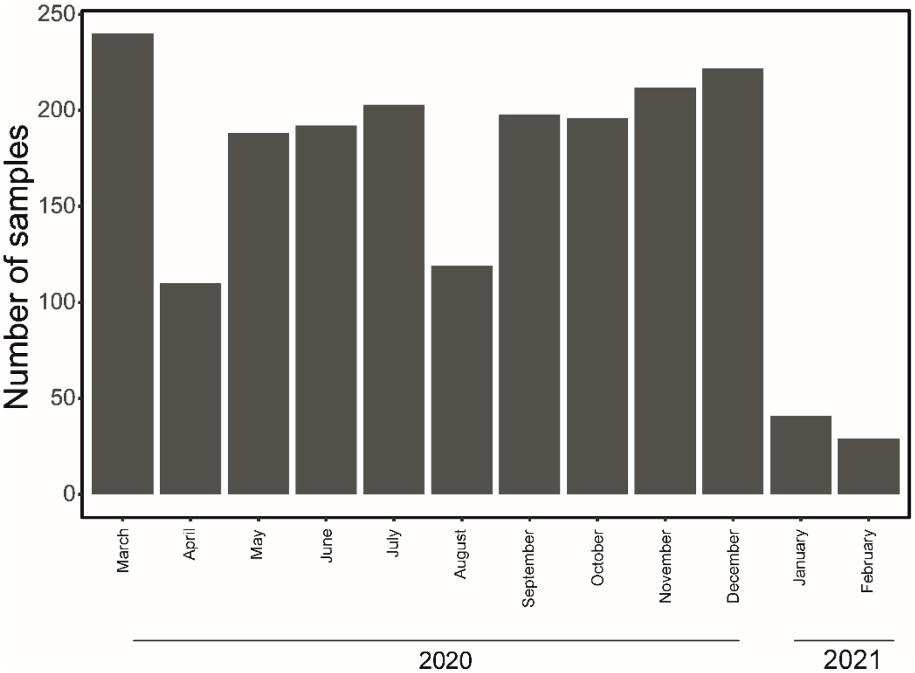

**Figure S4. Number of respiratory samples in the dataset per month.**

**Table S1. Sex distribution of the persons from which respiratory samples were gathered.**

| | Female | Male | Total |
|---|---|---|---|
| SARS-CoV-2 negative | 457 (49.0%) | 476 (51.0%) | 933 |
| SARS-CoV-2 positive | 489 (48.1%) | 528 (51.9%) | 1017 |

| | | | |
|---|---|---|---|
| Total | 946 (48.5%) | 1004 (51.5%) | 1950 |

***Section 2.2: Characteristics of the serum samples sent to the National Institute of Public Health Research (INSPI), Guayaquil, Ecuador 2020-2021***

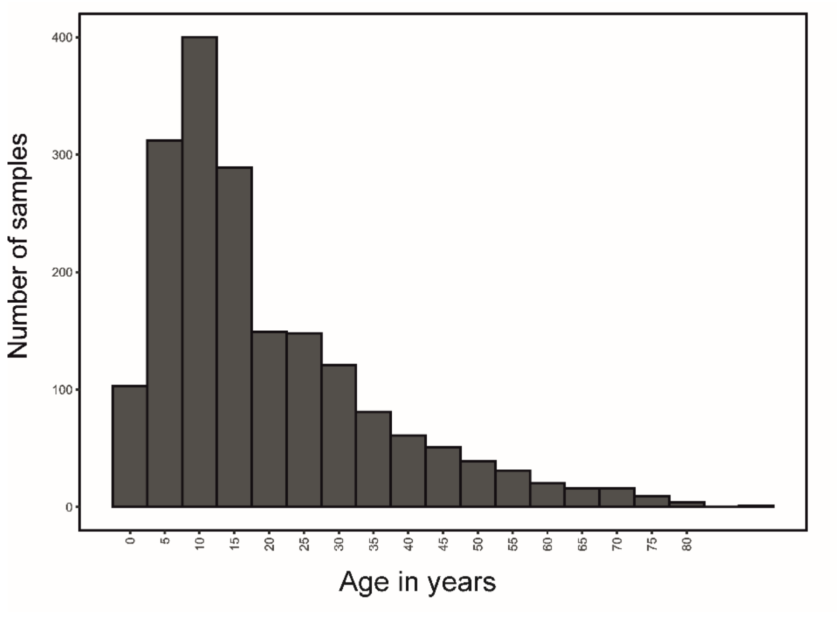

**Figure S5. Age distribution of the patients with fever of unknown origin from which serum samples were available.**

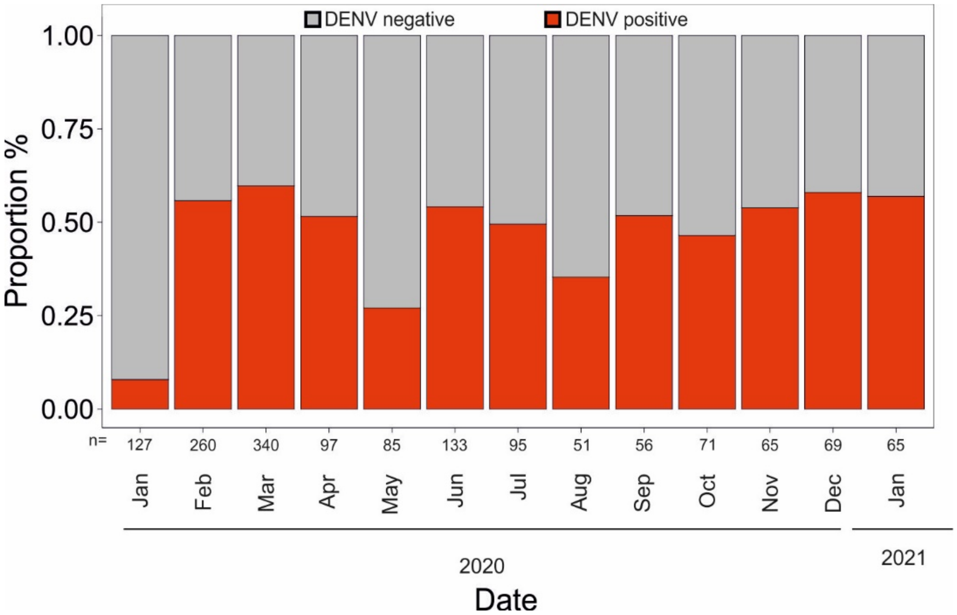

**Figure S6. Proportion of patients diagnosed with acute Dengue virus infection.**

**Table S2. Data from patients with fever of unknown origin investigated at INSPI**

| Characteristic | Overall, N = 1,967[1] | negative, N = 1,563[1] | positive, N = 404[1] |
|---|---|---|---|
| **Sex** | | | |
| Female | 947 (49%) | 749 (49%) | 198 (53%) |
| Male | 970 (51%) | 791 (51%) | 179 (47%) |
| Unknown | 50 | 23 | 27 |
| **Age*** | 14 (8, 27) | 15 (8, 28) | 12 (8, 24) |
| **Year** | | | |
| 2020 | 1,851 (94%) | 1,501 (96%) | 350 (87%) |
| 2021 | 116 (5.9%) | 62 (4.0%) | 54 (13%) |

| Characteristic | Overall, N = 1,967[1] | negative, N = 1,563[1] | positive, N = 404[1] |
|---|---|---|---|
| **Province** | | | |
| Azuay | 22 (1.1%) | 21 (1.4%) | 1 (0.3%) |
| Bolivar | 24 (1.3%) | 21 (1.4%) | 3 (0.8%) |
| Cañar | 10 (0.5%) | 4 (0.3%) | 6 (1.3%) |
| Carchi | 6 (0.3%) | 5 (0.3%) | 1 (0.3%) |
| Chimborazo | 12 (0.6%) | 12 (0.8%) | 0 (0%) |
| Cotopaxi | 6 (0.3%) | 6 (0.4%) | 0 (0%) |
| El Oro | 37 (1.9%) | 35 (2.3%) | 2 (0.5%) |
| Esmeraldas | 30 (1.6%) | 25 (1.6%) | 5 (1.3%) |
| Galapagos | 3 (0.2%) | 3 (0.2%) | 0 (0%) |
| Guayas | 997 (52%) | 728 (47%) | 269 (71%) |
| La Libertad | 1 (<0.1%) | 0 (0%) | 1 (0.3%) |
| Loja | 48 (2.5%) | 48 (3.1%) | 0 (0%) |
| Los Rios | 169 (8.8%) | 164 (11%) | 5 (1.3%) |
| Manabí | 258 (13%) | 221 (14%) | 37 (9.8%) |
| Morona Santiago | 15 (0.8%) | 9 (0.6%) | 6 (1.6%) |

| Characteristic | Overall, N = 1,967[1] | negative, N = 1,563[1] | positive, N = 404[1] |
|---|---|---|---|
| Napo | 20 (1.0%) | 19 (1.2%) | 1 (0.3%) |
| Orellana | 14 (0.7%) | 13 (0.8%) | 1 (0.3%) |
| Pastaza | 5 (0.3%) | 4 (0.3%) | 1 (0.3%) |
| Pichincha | 83 (4.3%) | 67 (4.4%) | 16 (4.2%) |
| Santa Elena | 7 (0.4%) | 4 (0.3%) | 3 (1.1%) |
| Santo Domingo | 92 (4.8%) | 84 (5.5%) | 8 (2.1%) |
| Sucumbíos | 36 (1.9%) | 30 (1.9%) | 6 (1.6%) |
| Zamora Chinchipe | 21 (1.1%) | 16 (1.0%) | 5 (1.3%) |
| N.A. | 51 | 24 | 27 |
| **Month** | | | |
| January-2020 | 145 (7.4%) | 145 (9.3%) | 0 (0%) |
| February-2020 | 285 (14%) | 285 (18%) | 0 (0%) |
| March-2020 | 473 (24%) | 473 (30%) | 0 (0%) |
| April-2020 | 99 (5.0%) | 85 (5.4%) | 14 (3.5%) |
| May-2020 | 160 (8.1%) | 115 (7.4%) | 45 (11%) |
| June-2020 | 149 (7.6%) | 95 (6.1%) | 54 (13%) |

| Characteristic | Overall, N = 1,967[1] | negative, N = 1,563[1] | positive, N = 404[1] |
|---|---|---|---|
| July-2020 | 111 (5.6%) | 65 (4.2%) | 46 (11%) |
| August-2020 | 80 (4.1%) | 40 (2.6%) | 40 (9.9%) |
| September-2020 | 82 (4.2%) | 51 (3.3%) | 31 (7.7%) |
| October-2020 | 89 (4.5%) | 48 (3.1%) | 41 (10%) |
| November-2020 | 83 (4.2%) | 45 (2.9%) | 38 (9.4%) |
| December-2020 | 95 (4.8%) | 54 (3.5%) | 41 (10%) |
| January-2021 | 83 (4.2%) | 46 (2.9%) | 37 (9.2%) |
| February-2021 | 33 (1.7%) | 16 (1.0%) | 17 (4.2%) |

[1]n (%); *Median (IQR); N.A.; Not available

**Section 3. Serologic testing**

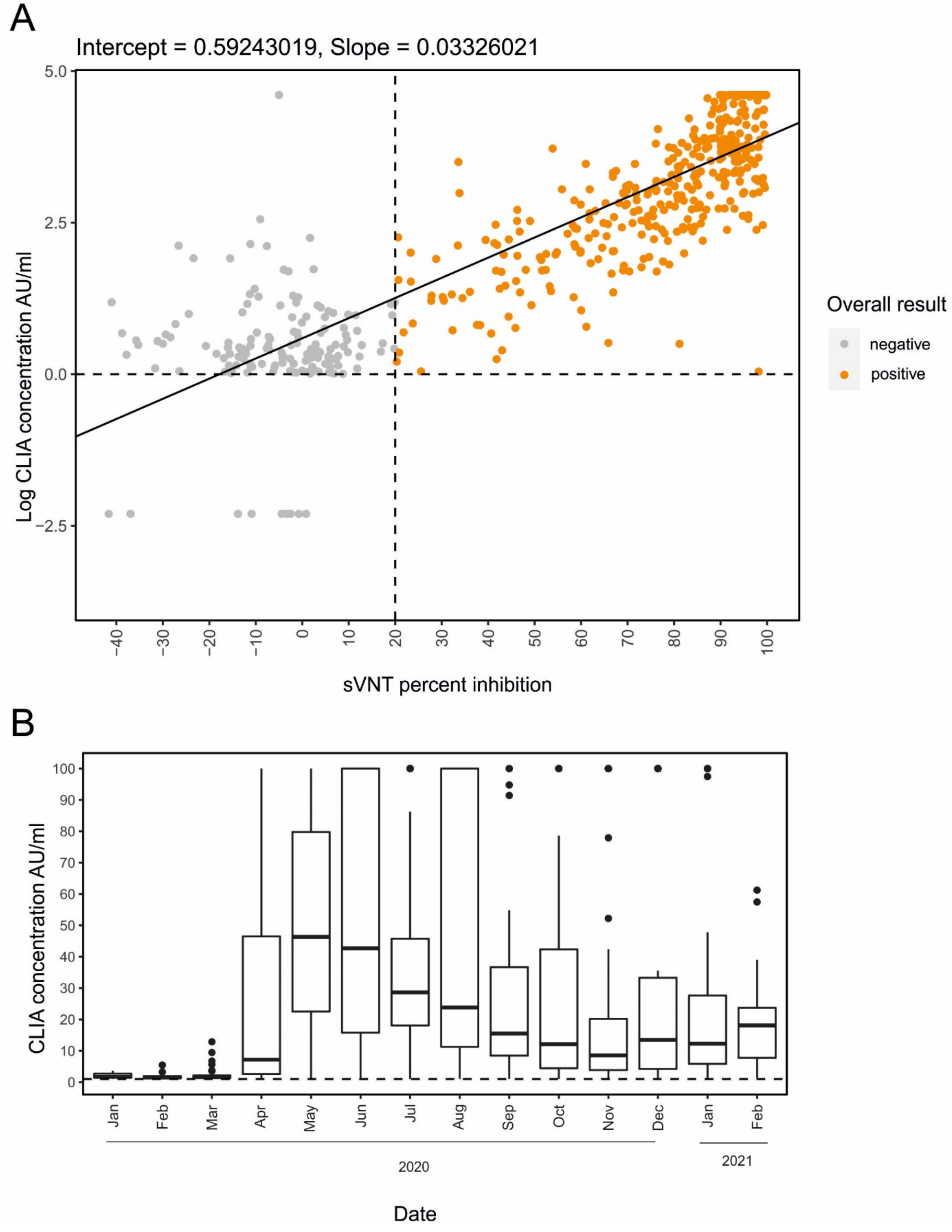

**Figure S7. Serologic testing.**

A) IgG antibody screening per patient. Only samples yielding positive results in a chemiluminescence immunoassay using the SARS-CoV-2 spike receptor-binding domain as antigen (CLIA; SARS-CoV-2 S-RBD IgG kit; Snibe Diagnostic, China) and a confirmatory SARS-CoV-2 surrogate virus neutralization test (sVNT; GenScript, USA) were considered positive. B) CLIA concentration across time in the tested samples. The line inside the Tukey boxplots denotes the median CLIA concentration per month. The length of the box is the interquartile range (IQR). Dots denote outliers. AU/ml: absorbance units per milliliter. Dotted line represents the limit where samples are considered positive as per manufacturer's instructions.

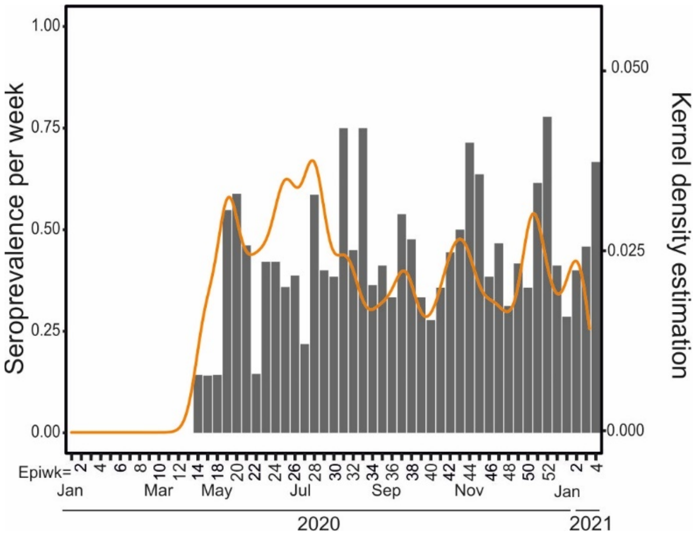

**Figure S8. Seroprevalence estimates using only samples from coastal Ecuador.**

Calculation of seroprevalence per epidemiological week using only serum samples of gathered only from coastal Ecuador.

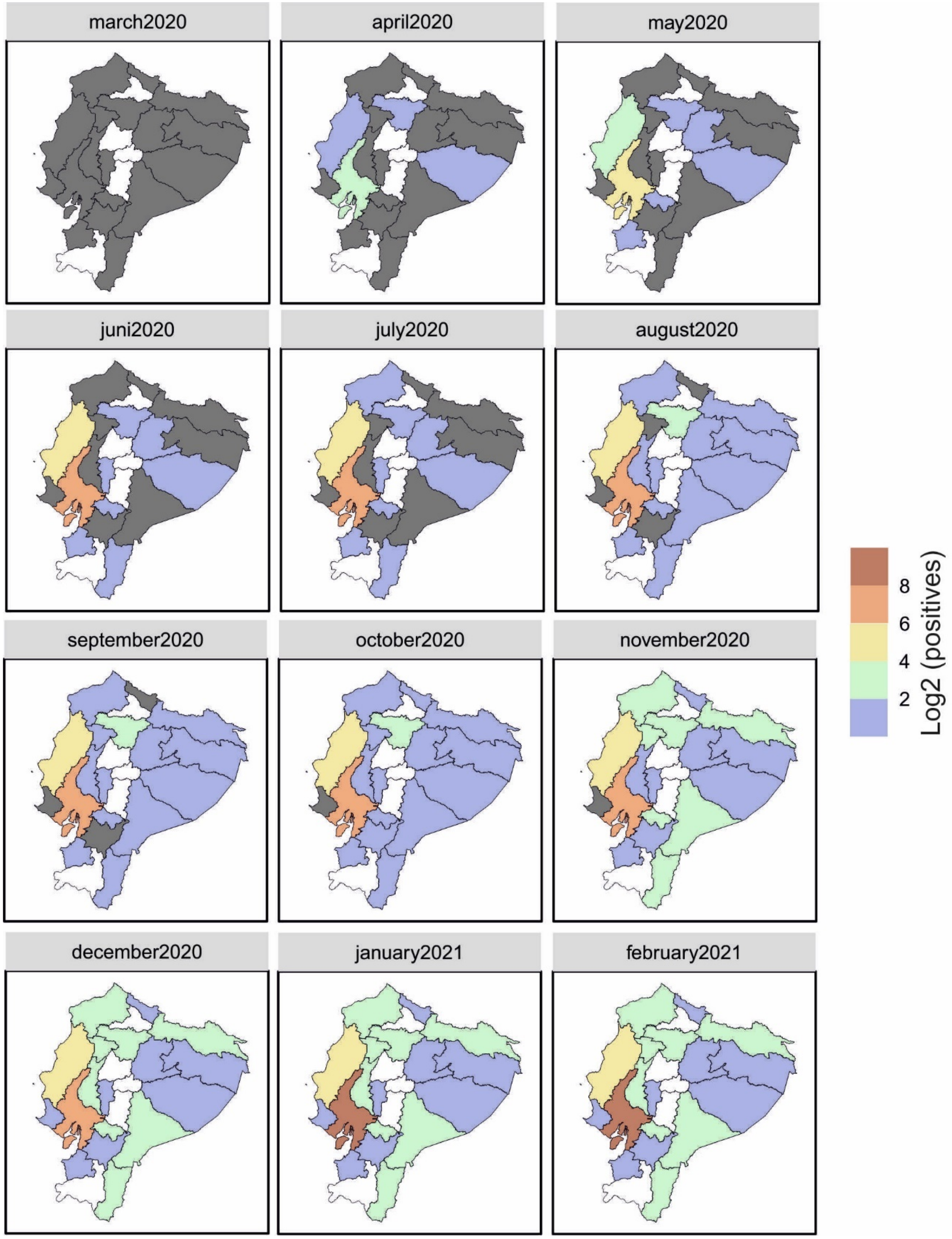

march2020
april2020
may2020
juni2020
july2020
august2020
september2020
october2020
november2020
december2020
january2021
february2021
8
6
4
2
Log2 (positives)

**Figure S9. Temporal distribution of patients with SARS-CoV-2-specific antibodies during the study period in Ecuador (IgG).**

Cumulative count of SARS-CoV-2 IgG antibody-positive patients per province per month is shown. Log scale was performed for clarity of presentation**.**

**Section 4. Non-pharmaceutical interventions in Ecuador.**

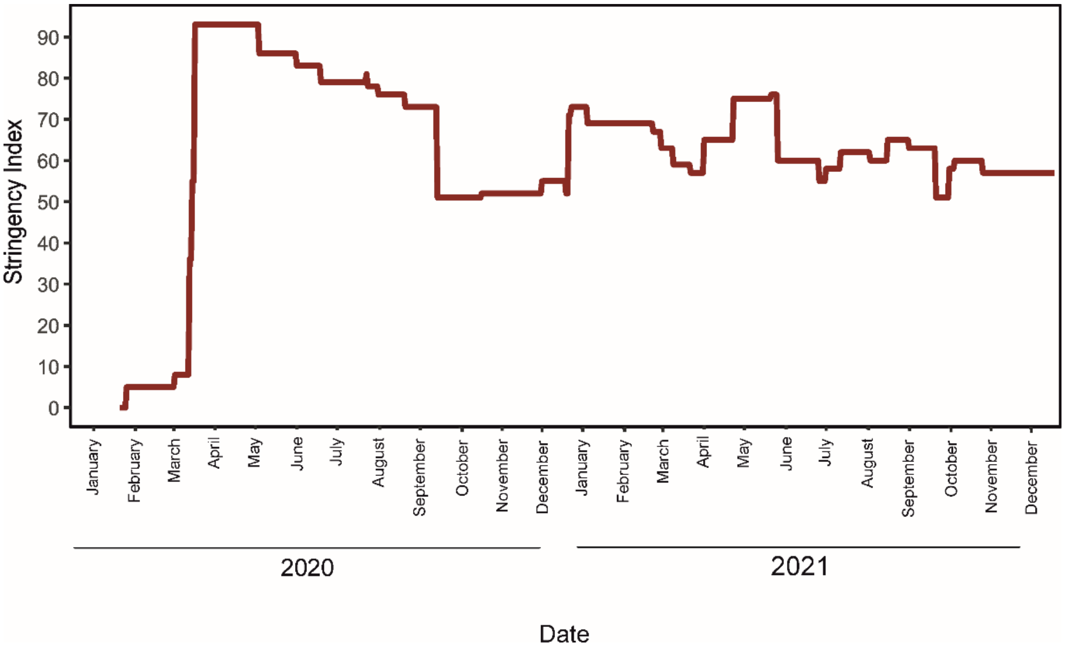

**Figure S10. OxCGRT stringency index in 2020-2021 per month in Ecuador.**

The OxCGRT stringency index was gathered from Our World in Data (Roser M. Coronavirus Pandemic (COVID-19). *Our World in Data* 2020).

**Section 5. Susceptible-exposed-infectious-recovered (SEIR) model data.**

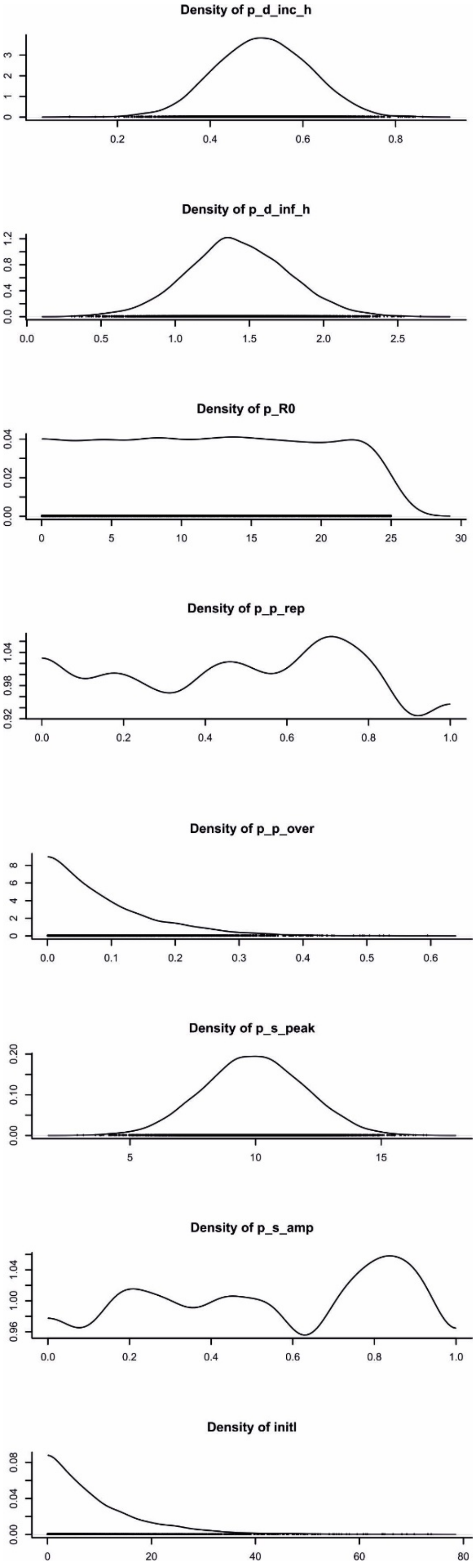

**Figure S11. SEIR model full prior probability distributions.**

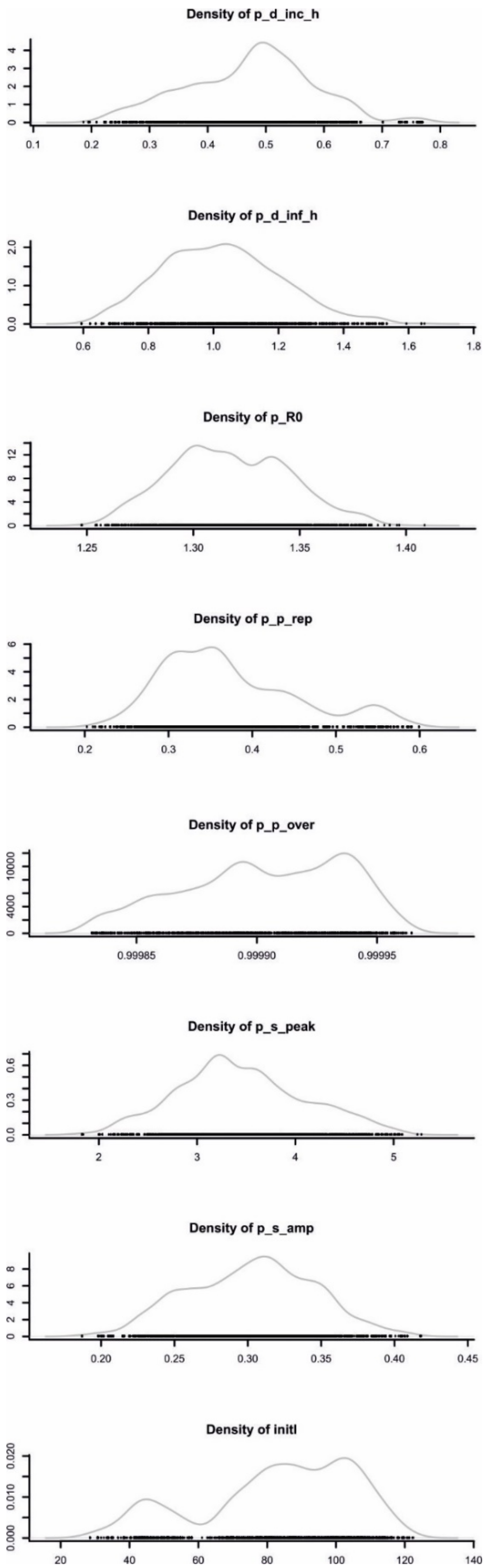
Density of p_d_inc_h
0 1 2 3 4
0.1 0.2 0.3 0.4 0.5 0.6 0.7 0.8
Density of p_d_inf_h
0.0 1.0 2.0
0.6 0.8 1.0 1.2 1.4 1.6 1.8
Density of p_R0
0 4 8 12
1.25 1.30 1.35 1.40
Density of p_p_rep
0 2 4 6
0.2 0.3 0.4 0.5 0.6
Density of p_p_over
0 4000 10000
0.99985 0.99990 0.99995
Density of p_s_peak
0.0 0.3 0.6
2 3 4 5
Density of p_s_amp
0 2 4 6 8
0.20 0.25 0.30 0.35 0.40 0.45
Density of initI
0.000 0.010 0.020
20 40 60 80 100 120 140

**Figure S12. SEIR model full posterior probability distributions.**

### Section 6. Molecular detection of SARS-CoV-2 and other respiratory viruses

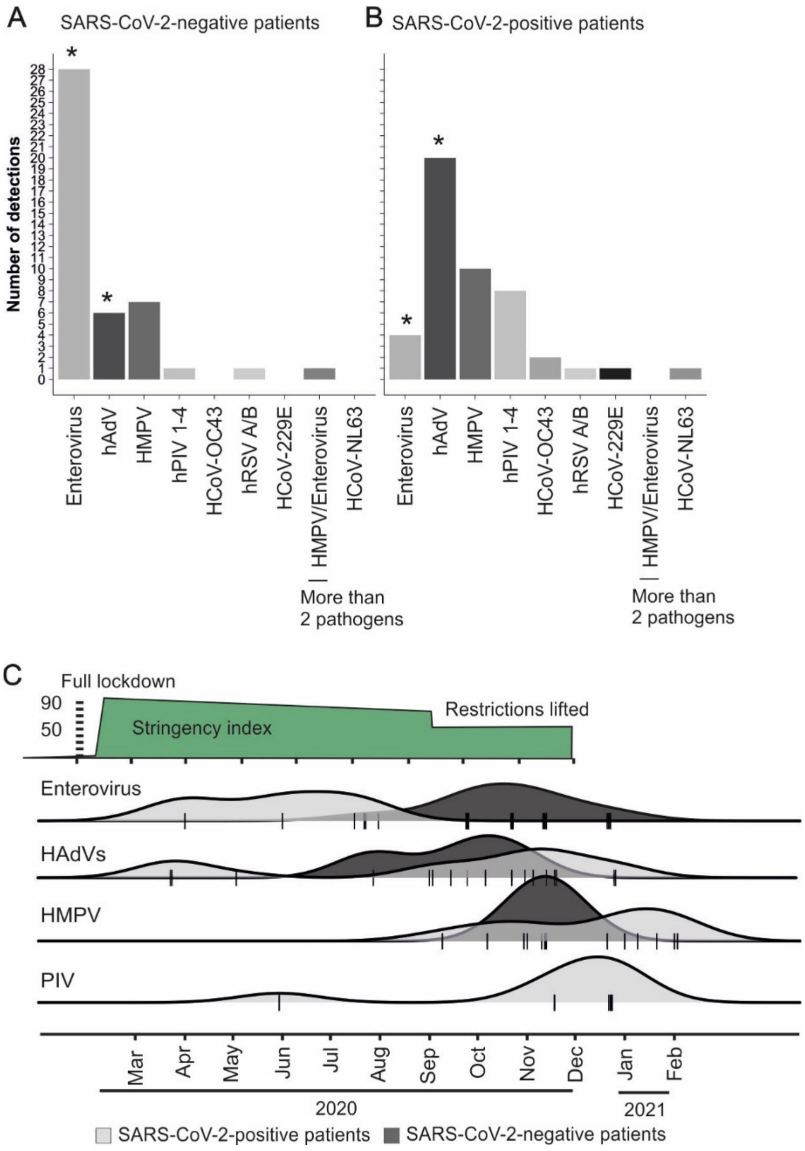

**Figure S13. Individual detection of respiratory viruses in SARS-CoV-2-positive and -negative patients.**

(A) and (B) Overall detection of other respiratory viruses. Common cold coronaviruses HCoV-OC43, -229E, -NL63, Adenovirus (HAdV), Respiratory syncytial viruses A and B (hRSV), metapneumovirus (hMPV), Enterovirus and Parainfluenza virus 1-4 (hPIV 1-4) in SARS-CoV-2-positive and -negative patients. Asterisk denotes statistically significant difference between SARS-CoV-2 confirmed- and negative-patients (Fisher's exact test; $p<0.01$). (B) Individual pathogen detection in SARS-CoV-2- confirmed and -negative patients. Abbreviations as in (A).

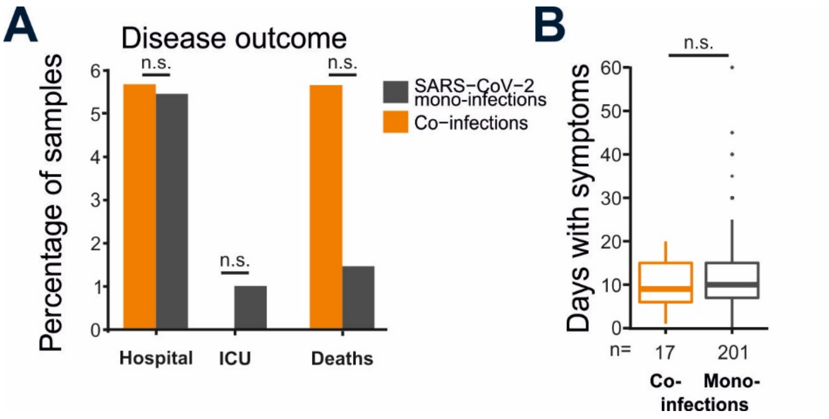

**Figure S14. Disease outcome, days with symptoms and symptomatology of SARS-CoV-2 co-infected versus mono-infected patients.**

(A) Percentage of co-infected and SARS-CoV-2 mono-infected patients that were hospitalized, admitted to the intensive care unit (ICU) and reported mortality. N.s. not significant. Right: Boxplots showing the mean days with symptoms of co-infected and SARS-CoV-2 mono-infected patients (t-test with Bonferroni correction.; p =0.08). Dots represent outliers. (B) Percentage of recorded days with symptoms of SARS-CoV-2 co-infected versus mono-infected patients. N.s. not significant.

**Section 7. Phylogenetic analyses of SARS-CoV-2**

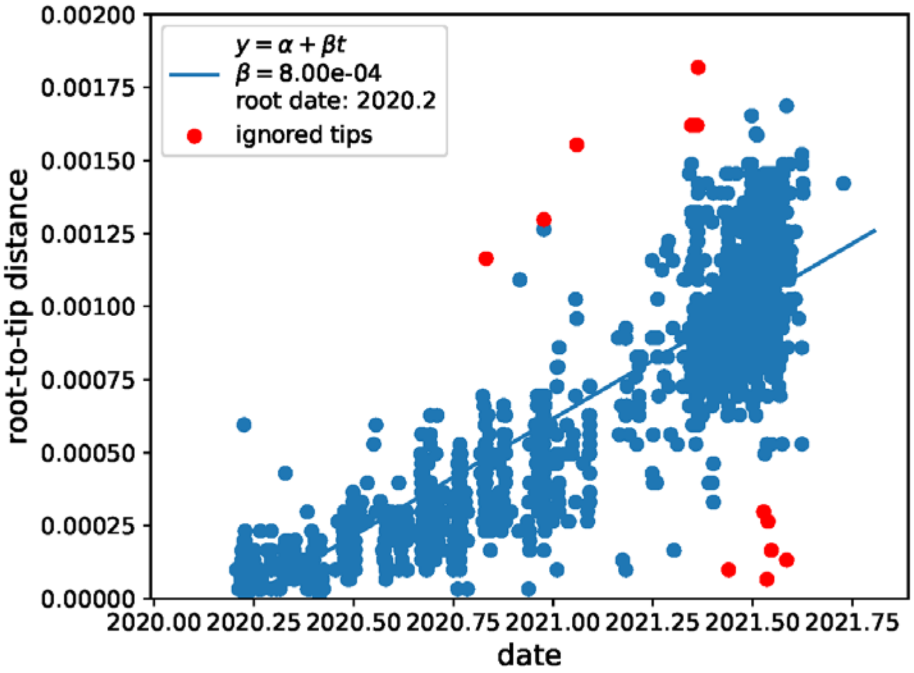

**Figure S15. Root-to-tip distance of Ecuadorian SARS-CoV-2 sequences.**

Some tips are automatically ignored by the Treetime program to minimize variance of root-to-tip distances.

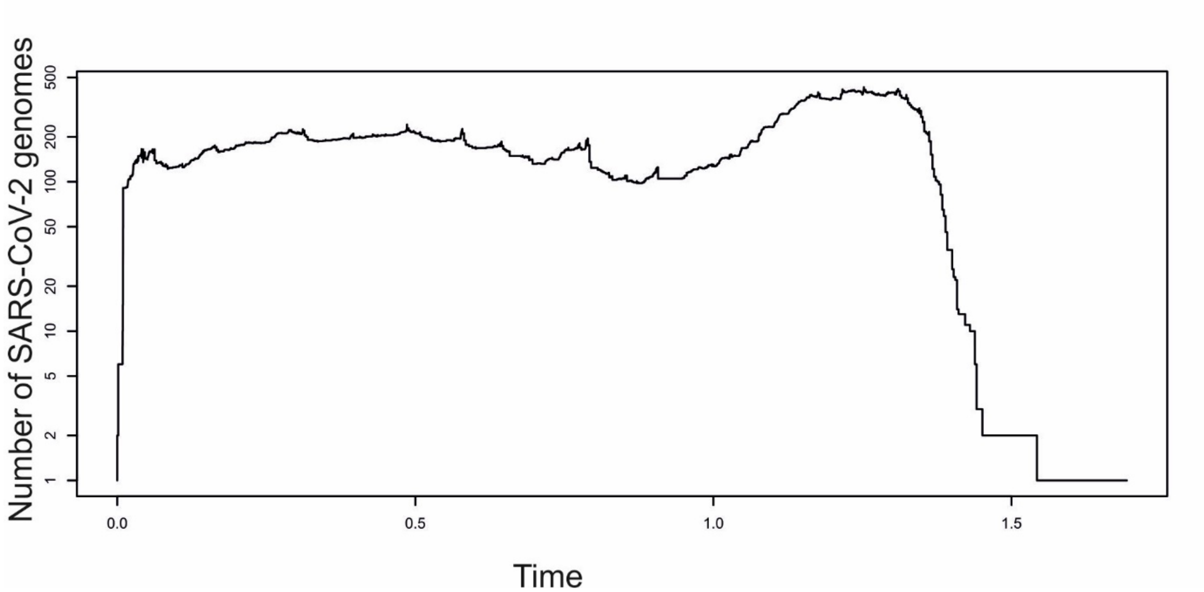

**Figure S16. Number of SARS-CoV-2 tips per time in the time-stamped phylogenetic tree.**

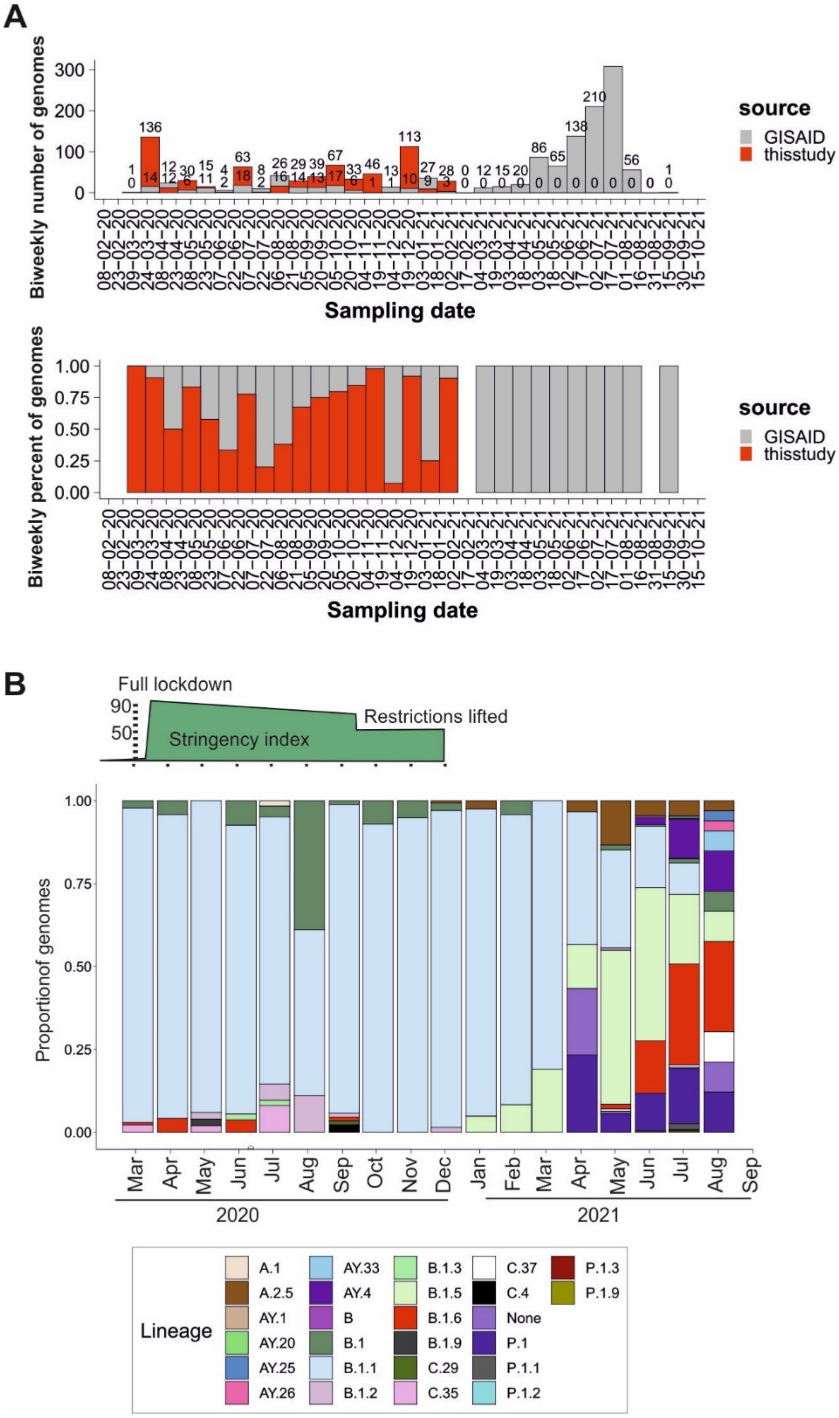

**Figure S17. Summary of SARS-CoV-2 lineages in Ecuador.**

(A) Number and proportion of SARS-CoV-2 genomes used for this study. Red denotes newly generated genomes for this study overtime. (B) Proportion of genomes from different lineages in the dataset.

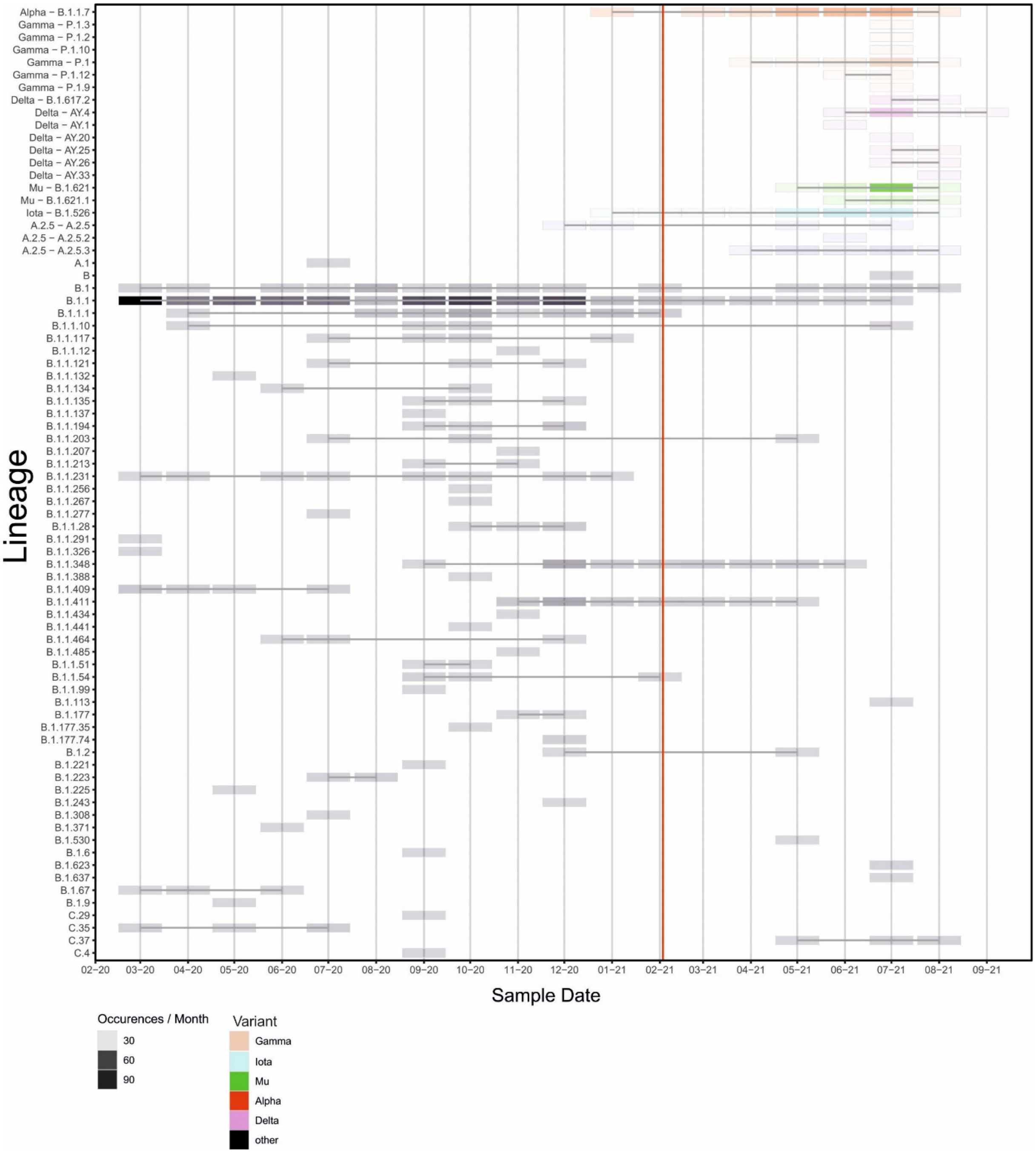

**Figure S18. Detail of SARS-CoV-2 lineages in Ecuador.**

Individual detection of lineages in the dataset. Red line denotes the date from which afterwards only GISAID sequences were available. Colors represent the former variants of concern, based on World Health Organization classification.

### Section 8. GISAID sequence metadata

**Table S3. Metadata of GISAID sequences used for this study.**

Data Availability GISAID Identifier: EPI_SET_20220712gt doi: 10.55876/gis8.220712gt All genome sequences and associated metadata in this dataset are published in GISAID's EpiCoV database. To view the contributors of each individual sequence with details such as accession number, Virus name, Collection date, Originating Lab and Submitting Lab and the list of Authors, visit 10.55876/gis8.220712gt. EPI_SET_20220712gt is composed of 973 individual genome sequences. All sequences in this dataset are compared relative to hCoV-19/Wuhan/WIV04/2019 (WIV04), the official reference sequence employed by GISAID (EPI_ISL_402124). Learn more at https://gisaid.org/WIV04.
